# Impaired Neutralizing Antibody Response to COVID-19 mRNA Vaccines in Cancer Patients

**DOI:** 10.1101/2021.10.20.21265273

**Authors:** Cong Zeng, John P. Evans, Sarah Reisinger, Jennifer Woyach, Christina Liscynesky, Zeinab El Boghdadly, Mark P. Rubinstein, Karthik Chakravarthy, Linda Saif, Eugene M. Oltz, Richard J. Gumina, Peter G. Shields, Zihai Li, Shan-Lu Liu

## Abstract

There is currently a critical need to determine the efficacy of SARS-CoV-2 vaccination for immunocompromised patients. In this study, we determined the neutralizing antibody response in 160 cancer patients diagnosed with chronic lymphocytic leukemia (CLL), lung cancer, breast cancer, and various non-Hodgkin’s lymphomas (NHL), after they received two doses of mRNA vaccines. Serum from 46 mRNA vaccinated health care workers (HCWs) served as healthy controls. We discovered that (1) cancer patients exhibited reduced neutralizing antibody titer (NT_50_) compared to HCWs; (2) CLL and NHL patients exhibited the lowest NT_50_ levels, with 50-60% of them below the detection limit; (3) mean NT_50_ levels in patients with CLL and NHL was ∼2.6 fold lower than those with solid tumors; and (4) cancer patients who received anti-B cell therapy exhibited significantly reduced NT_50_ levels. Our results demonstrate an urgent need for novel immunization strategies for cancer patients against SARS-CoV-2, particularly those with hematological cancers and those on anti-B cell therapies.

## Main

In response to the global public health crisis caused by the COVID-19 pandemic, several SARS-CoV-2 vaccines were rapidly developed including the Pfizer/BioNTech BNT162b2 and Moderna mRNA-1273 mRNA vaccines. However, clinical trials of these mRNA vaccines did not investigate their efficacy in vulnerable populations, including immunocompromised patients. With rising vaccination rates and an easing of public health measures, there is a critical need to determine the efficacy of SARS-CoV-2 vaccination for such patients, who may experience a reduced efficacy of administered vaccines^1^. It has already been demonstrated that organ transplant recipients, who are under immunosuppressive therapy to prevent rejection, exhibit reduced responsiveness to SARS-CoV-2 vaccination^2,3^. Cancer patients represent another critical population of immunocompromised individuals who, due to the nature of the disease or to treatment with immunomodulatory therapies, may not exhibit a robust response to mRNA vaccination. A better understanding of the factors governing response to vaccination in cancer patients is critical to inform clinical decisions about the need for booster doses, the timing of vaccine administration, the need to interrupt treatment courses for vaccination, and general guidance about the level of protection achieved by vaccination in cancer patients. To this end, this study examines the neutralizing antibody response to Pfizer/BioNTech BNT162b2 and Moderna mRNA-1273 vaccination in a cohort of patients with solid tumor and hematological malignancies.

The study population included 160 cancer patients (54 chronic lymphocytic leukemia (CLL), 45 non-Hodgkin’s lymphoma (NHL), 29 lung cancer, 30 breast cancer, and 2 breast cancer with CLL) recruited through medical record screening for vaccine appointments or recent post-vaccine administration, as well as an independent cohort of 46 health care workers (HCWs), who have no history of cancer. Cancer patients had a median age of 66 years while the median age of HCWs were 38 years. No cancer patient or HCW was COVID-19 positive as confirmed by nucleocapsid-based ELISA. About 61% of cancer patients (n=98) and 52% of the HCWs (n=24) received BNT162b2, compared to 39% (n=62) and 48% (N=22) who received the mRNA-1273, respectively. We collected serum samples for 159/160 cancer patients between 31 and 232 days (median 134 days) post-second dose, and HCW serum samples were obtained at 6 months post-second dose. Cancer diagnoses and treatments of the patients are shown in **Table 1**. The largest treatment groups were 47 patients with B-cell malignancies (28 CLL and 19 NHL) who received B cell depletion therapy or other B cell-suppressing drugs (such as anti-CD20 monoclonal antibodies and Bruton tyrosine kinase (BTK) inhibitors) during the study period; and 46% (n=28) of solid tumor patients received immune checkpoint inhibitors against PD-1 or PD-L1.

**Table 1.**
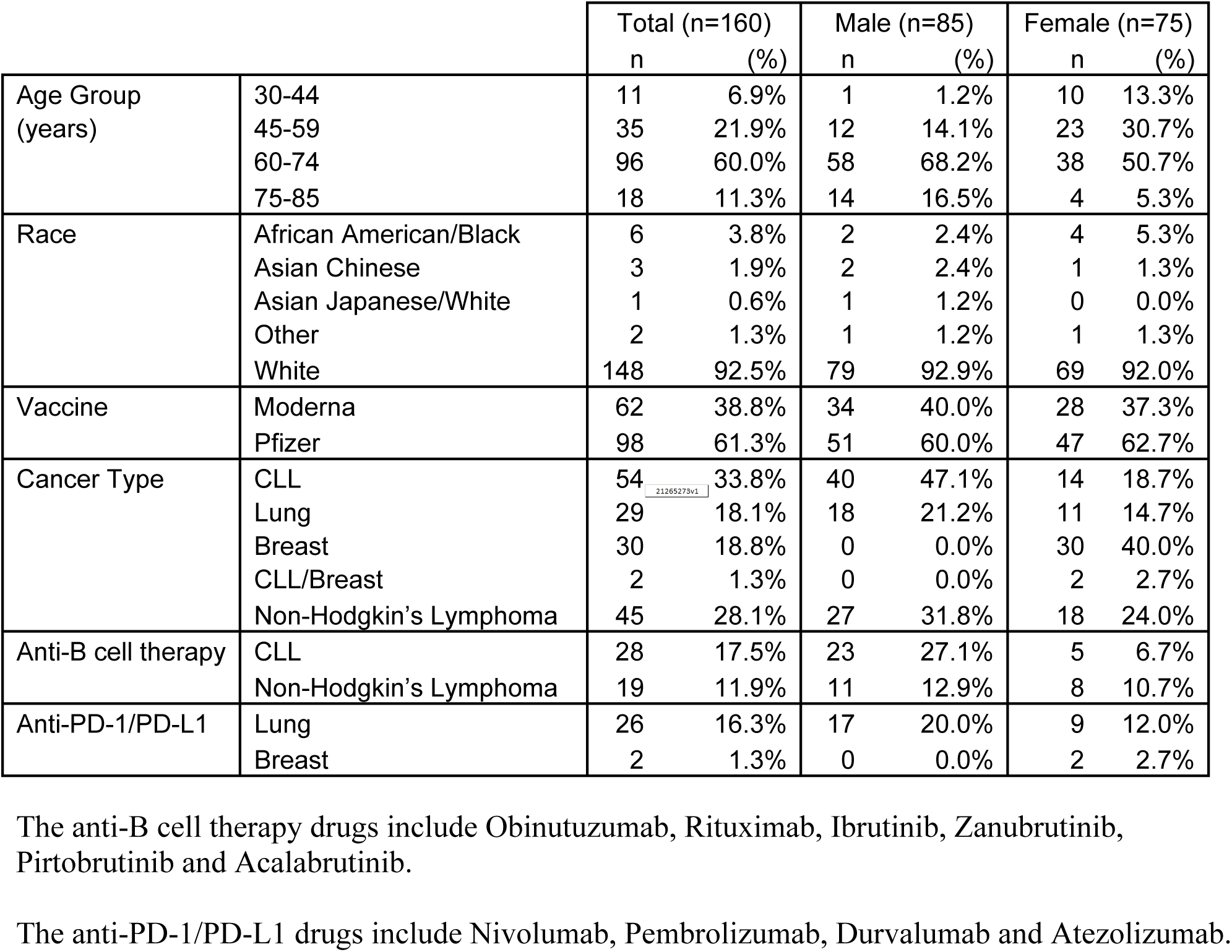
Demographic information of cancer patients.

We assessed sera for neutralizing antibody titers using a secreted *Gaussia*-luciferase SARS-CoV-2-pseudotyped-lentivirus-based virus neutralization assay as previously described^4^. Briefly, pseudotyped virus was incubated with serial dilutions of patient sera and used to infect HEK293T-ACE2 cells (BEI NR-52511). Infected cells then secreted *Gaussia*-luciferase into the culture media which was harvested 48hrs and 72hrs after infection, and luminescence was measured by a BioTek Cytation5 plate-reader. The resulting luciferase output was used to calculate a neutralization titer at 50% efficiency of maximal inhibition (NT_50_). To ensure valid comparisons, the serum samples of all cancer patients and HCWs were processed side-by-side in the same experiment.

We first compared the neutralizing antibody titers of cancer patients with those of HCWs. Overall, cancer patients exhibited reduced neutralizing antibody responses, with a mean NT_50_ of 220 compared to a mean NT_50_ of 522 for HCWs (**Fig. 1a**); this is despite the relatively shorter median time (134 days) after the second dose of vaccination for cancer patients as compared to HCW, which is an average of ∼180 days. Patients with CLL exhibited the lowest neutralizing antibody response, with over 61% (n=34) of patients exhibiting undetectable NT_50_ values (below 40), compared to 49%, 31%, and 28% for NHL (n=22), lung cancer (n=9), and breast cancer patients (n=9), respectively (**Fig. 1b**). The mean NT_50_ of patients with CLL and NHL (158 and 127, respectively) was ∼2.6 fold lower than that of solid tumor patients (369) (**Fig. 1a**). This is consistent with reports that SARS-CoV-2 infection induced weak humoral immune responses in patients with hematological cancers^5^. Interestingly, there were a few CLL patients that exhibited high titer while none were observed for the NHL patients (**Fig. 1b**).

**Figure 1.**
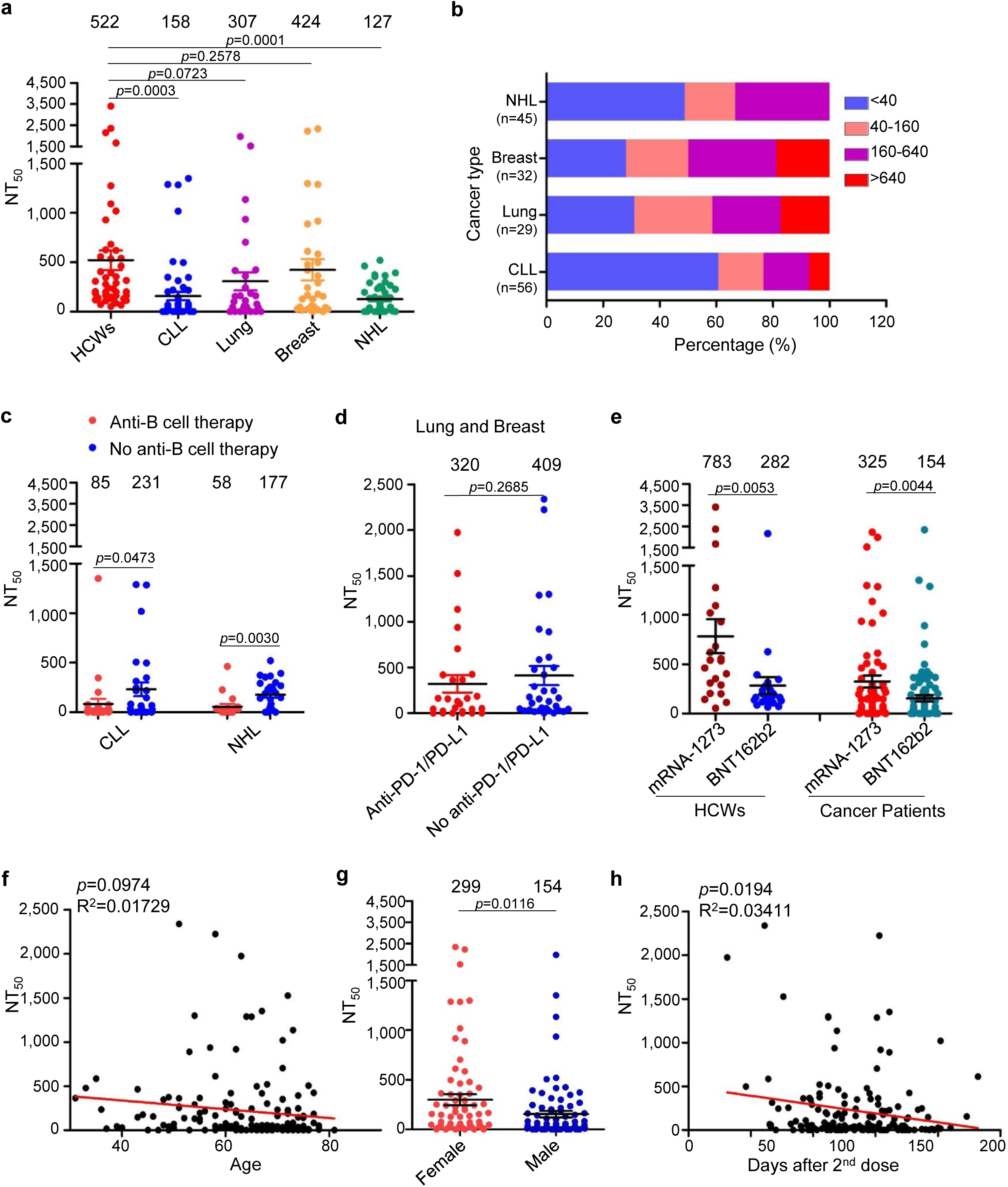
Neutralization of SARS-CoV-2 spike-pseudotyped lentivirus by sera of cancer patients and health care workers. (**a**) Comparison of 50% neutralization titer (NT_50_) between cancer patients and health care workers (HCWs). Serially diluted sera were incubated with SARS-CoV-2 spike-pseudotyped lentivirus, followed by infection of 293T-ACE2 cells. The assay was carried out side by side for samples of healthy individuals and cancer patients to ensure valid comparisons. (**b**) Distribution ranges of NT_50_ among four cancer patient groups. Note that 2 patients who had both CLL and breast cancer were included in each group. (**c**) Comparison of NT_50_ between anti-B cell therapy and no anti-B cell therapy in cancer patients. Twenty-eight out of the 54 CLL patients and 19 out of the 45 NHL patients received anti-B cell therapy, with drugs including BTK inhibitors and anti-CD20 monoclonal antibodies. (**d**) Comparison of NT_50_ between Anti-PD1/PD-L1 and no anti-PD1/PD-L1 treatment in lung and breast cancer patients. (**e**) Comparison of NT_50_ between Moderna and Pfizer vaccinees in health care workers (HCWs) and cancer patients. (**f**) Correlative analysis between NT_50_ values and ages of cancer patients. (**g**) Comparison of NT_50_ values between male and female cancer patients. (**h**) Correlative analysis between NT_50_ values and days of collection after the second dose of vaccination. All correlative analyses were performed using Prism 5 (f and h). In all cases, NT_50_ values indicated at top were calculated by taking the inverse of the 50% inhibitory dilution values obtained from least squares regression non-linear curve modeled with Prism. Statistical significance was determined by a one-tailed unpaired t-test. CLL: Chronic Lymphocytic Leukemia; NHL: Non-Hodgkin’s Lymphoma.

Given the common usage of B-cell depleting therapies in the treatment of hematological cancers^6-8^ and their likelihood of impacting vaccine efficacy, we then examined the effect of anti-B-cell therapy on neutralizing antibody response. The treatment included anti-CD20 antibodies Obinutuzumab and Rituximab, as well as BTK inhibitors Ibrutinib, Zanubrutinib, Pirtobrutinib, and Acalabrutinib. Notably, we found that CLL and NHL patients who received anti-B cell therapy exhibited 2.7-fold (p = 0.0483) and 3.1-fold (p = 0.0030) reduced neutralizing antibody response to mRNA vaccine compared to those without anti-B cell therapy, respectively (**Fig. 1c**).

The programmed death-1 (PD-1) receptor is an important immune checkpoint molecule that promotes exhaustion/dysfunction in chronically activated T-cells^9^. Disruption of PD-1 or its ligand PD-L1 is a common treatment to rejuvenate T cell function in cancer patients^10,11^. Given this role, we examined how anti-PD-1/PD-L1 treatments might modulate the host immune response to mRNA vaccination. However, we did not find significant differences in NT_50_ or development of immune-related adverse events between anti-PD-1/PD-L1 antibody treated and un-treated lung/breast cancer patients (**Fig. 1d**).

Other factors potentially impacting immune stimulation were also assessed, including age and gender of patients, types of vaccine received and time of sample collection. Moderna mRNA-1273 outperformed Pfizer BNT162b2 vaccine in mean NT_50_ by 2.8-fold for HCWs (p = 0.0053) and 2.1-fold for cancer patients (p = 0.0044) (**Fig. 1e**). This is consistent with our previous findings that Moderna mRNA-1273 vaccinated individuals exhibit higher NT_50_ levels compared to Pfizer BNT162b2^12^. Given previous findings that neutralizing antibody response to mRNA vaccination is age dependent^13^, we also examined the possible correlation between age and NT_50_ titer. However, no significant correlation between age and NT_50_ values was observed in these cancer patients (**Fig. 1f**). Notably, while male patients have been shown to exhibit higher NT_50_ levels following COVID-19 disease^14^, we found here that female patients in fact exhibited a higher level of virus neutralization with a mean NT_50_ of 299 compared to 154 for males (p = 0.0116; **Fig. 1g**). This likely reflects an overrepresentation of older patients and patients with hematological cancers in males in our cohort (**Table 1**).

Given increasing concerns about declining efficacy of SARS-CoV-2 vaccines^15,16^, we also examined the correlation between NT_50_ and time post second vaccine dose for these cancer patients. Indeed, we observed a significant, negative correlation (p = 0.0194) between time after second dose of mRNA vaccination and NT_50_ value (**Fig. 1h**). These results confirm the waning immune protection of neutralizing antibodies that are conferred by mRNA vaccination.

In summary, by using a sensitive high-throughput lentivirus-based SARS-CoV-2 neutralization assay^4^, we have examined the neutralizing antibody response of 160 cancer patients and compared, side by side, with that of 46 healthy HCWs. We observed about an approximately 2.4-fold lower neutralizing antibody response in the cancer patients as compared to HCWs, following Pfizer/BioNTech BNT162b2 or Moderna mRNA-1273 vaccination, clearly demonstrating a reduced efficacy of SARS-CoV-2 spike antibody production among cancer patients. This, along with similar observations of some recent complementary studies^17-19^, should inform the development of novel immunization strategies for cancer patients. In particular, we find that patients with hematological cancers, such as CLL and NHL, are least likely to respond to mRNA vaccination, with 50-60% of these patients showing no detectable levels of neutralizing antibody against the SARS-CoV-2 spike. Given these findings, booster vaccines may be of particular importance for these groups of cancer patients, with some studies already underway^20^. Additionally, our finding that B cell depletion or suppression drug treatment significantly reduced the neutralizing antibody response to mRNA vaccines may indicate a need for immunization to occur during disruptions or suspensions in specific treatment protocols.

Finally, to better protect immunocompromised populations with increased risk to COVID-19, we must further investigate the duration of vaccine induced immunity as well as the efficacy of booster vaccine doses to determine how to maintain protective immunity in this patient population. Additionally, further study on quality and durability of antigen-specific T and memory B cell responses will provide a more comprehensive understanding of the immune response to SARS-CoV-2 vaccination in these immunocompromised groups. It is also critical to determine the impact of specific treatment protocols on vaccine induced immunity and immunity duration to better inform clinical decisions about the time of vaccination or boosting and the potential need for disruptions in treatment protocols. Results from this work provide critical virological and immunological information for protecting vulnerable populations.

## Data Availability

All data produced in the present study are available upon reasonable request to the authors

## Acknowledgments

We thank members of Liu lab for sharing reagents and discussion. We also thank Donna Bucci, Jamie Hamon, Chelsea Bolyard, Kevin Weller, and Taylor Chatlos from the Immune Monitoring and Discovery Platform of the Pelotonia Institute for Immuno-Oncology for assistance in sample collection and discussion. We thank the Clinical Research Center/Center for Clinical Research Management of The Ohio State University Wexner Medical Center and The Ohio State University College of Medicine in Columbus, OH, specifically, Francesca Madiai, Claire Carlin, Dina McGowan, Breona Edwards, and Trina Wemlinger, for collection, processing, and management of samples through the Cardiovascular Medicine JB Biorepository. We thank the NIH AIDS Reagent Program and BEI Resources for supplying important reagents that made this work possible. This work was supported by the Ohio State University Comprehensive Cancer Center and a fund provided by an anonymous private donor to The Ohio State University and NCI U54CA260582 to S.-L.L. The content is solely the authors’ responsibility and does not necessarily represent the official views of the NIH. S.-L.L. was additionally supported in part by NIH R01 AI150473. Z.L. and P.G.S were supported by multiple NIH grants. R.J.G. was supported by NIH R01 HL127442-01A1, NCI U54CA260582, and the Cardiovascular Medicine JB Biorepository, and the Robert J. Anthony Fund for Cardiovascular Research. L.S. and E.M.O. were also supported by NCI U54CA260582.

## Materials and Methods

### Vaccinated Cancer Patients and Health Care Workers

De-identified vaccinated health care worker (HCW)’s serum samples were collected under approved IRB protocols (2020H0228 and 2020H0527). Serum was collected 6 months after the second dose of Pfizer (n=24) and Moderna (n=22) SARS-CoV-2 mRNA vaccine. The ages of the vaccinated groups ranged from 26 years to 61 years (mean age=38.5).

Cancer patient serum samples were collected under an approved IRB protocol (2021C0041). Serum was collected 31∼232 days (median 134 days) after the second dose of Pfizer (n=98) and Moderna (n=62) SARS-CoV-2 mRNA vaccines. The age of vaccinated cancer patients ranged from 31 years to 81 years (mean age=66). These cancer patients consisted of 29 lung cancer, 54 chronic lymphocytic leukemia (CLL), 30 breast cancer, 2 CLL and breast cancer, and 45 various non-Hodgkin’s lymphomas. Patients were 47% (n=75) female and 53% (n=85) male. Twenty-eight of the 54 CLL patients and 19 of the 45 non-Hodgkin’s lymphomas patients received anti-B cell therapy, with drugs including BTK inhibitors and anti-CD20 mAbs during the study period. Twenty-six of the 29 lung cancer patients and 2 of the 30 breast cancer patients received anti-PD-1/PD-L1 therapy.

### Cell Culture

HEK293T cells (ATCC CRL-11268, CVCL_1926) and HEK293T-hACE2 cells (BEI NR-52511) were grown in DMEM (Gibco, 11965-092) supplemented with 10% (vol/vol) fetal bovine serum (Sigma, F1051) and 1% penicillin/streptomycin (HyClone, SV30010). Both cell lines were maintained at 37°C, and 5% CO2.

### Constructs

The construct used for the production of lentiviral pseudotypes was HIV-1 NL4.3-inGluc^1-3^, which was originally obtained from David Derse’s lab at NIH (National Cancer Institute, Frederick, Maryland, USA) and Marc Johnson’s lab at the University of Missouri (Columbia, Missouri, USA). This construct is based on a ΔEnv pNL4.3 HIV-1 vector and contains an anti-sense Gaussia luciferase (Gluc) gene with a sense intron. Gluc is secreted in mammalian cell culture^4^, and the intron and anti-sense orientation of the Gluc gene prevents the production of Gluc in the virus producer cells^1-3^. pcDNA3.1-SARS-CoV2-S-C9 encoding SARS-CoV-2 full-length spike was obtained from Fang Li’s lab at the University of Minnesota (St. Paul, Minnesota, USA).

### Virus Production

Lentiviral pseudotyped virus was produced as previously described^1^. Briefly, HEK293T cells were transfected with HIV-1 NL4.3-inGluc and pcDNA3.1-SARS-CoV2-S-C9 constructs in a 2:1 ratio using polyelthylenimine (PEI). Supernatants were harvested 24 hr, 48 hr, and 72 hr post-transfection and were pooled, aliquoted, and stored at −80°C.

### Pseudotype Virus Neutralization Assays

Pseudotyped virus neutralization assays (VNAs) were performed as previously described^1^. Briefly, cancer patients or vaccinee individual serum was 4-fold serially diluted in 96-well plate (Cellstar, 655180), resulting in a final volume of 60 μL. Subsequently, 100 μL of pseudotyped virus was added to the plate resulting in a final set of dilutions of 1:40, 1:160, 1:640, 1:2560, 1:10240, and no serum. Virus and serum mixture were incubated for 1 hr at 37°C, and then added to HEK293T-ACE2 cells seeded at 2 × 10^4^ cells/well. Media was changed after 6 hrs post-infection. At 48 hrs and 72 hrs after infection, 20 μL of media was collected from the cells and transferred to a white, flat-bottomed, polystyrene 96-well plate (Thermo Scientific, 236108). 20 μL of Gaussia luciferase substrate (0.1 M Tris (MilliporeSigma, #T6066) pH 7.4, 0.3 M sodium ascorbate (Spectrum, S1349), 10 μM coelenterazine (GoldBio, CZ2.5)) was added to the media and immediately read by a BioTek Cytation5 plate-reader.

### Quantification and Statistical Analysis

Data were analyzed as mean with Standard Error of Mean (SEM), Statistical analyses were performed using GraphPad Prism 5.0 as follows: One-way Analysis of Variance (ANOVA) with Bonferroni’s post-tests was used to compute statistical significance (p values) between multiple groups for multiple comparison or t-test was used for two groups for single comparison. The 50% neutralization titer (NT50) was determined using the half-maximal inhibitory concentration values of plasma samples, normalized to control infection, from their serial dilutions. NT50 values were calculated from VNA output using a non-linear regression with least-squares fit in GraphPad Prism5 (GraphPad Software, San Diego, California USA, www.graphpad.com).

## References

1. Romano, E., Pascolo, S. & Ott, P. Implications of mRNA-based SARS-CoV-2 vaccination for cancer patients. Journal for Immunotherapy of Cancer 9(2021).

2. Boyarsky, B.J., et al. Immunogenicity of a single dose of SARS-CoV-2 messenger RNA vaccine in solid organ transplant recipients. Jama 325, 1784–1786 (2021).

3. Rabinowich, L., et al. Low immunogenicity to SARS-CoV-2 vaccination among liver transplant recipients. Journal of Hepatology (2021).

4. Zeng, C., et al. Neutralizing antibody against SARS-CoV-2 spike in COVID-19 patients, health care workers, and convalescent plasma donors. JCI insight 5(2020).

5. Abdul-Jawad, S., et al. Acute immune signatures and their legacies in severe acute respiratory syndrome coronavirus-2 infected cancer patients. Cancer Cell 39, 257–275. e256 (2021).

6. Salles, G., et al. Rituximab in B-cell hematologic malignancies: a review of 20 years of clinical experience. Advances in therapy 34, 2232–2273 (2017).

7. Mössner, E., et al. Increasing the efficacy of CD20 antibody therapy through the engineering of a new type II anti-CD20 antibody with enhanced direct and immune effector cell–mediated B-cell cytotoxicity. Blood, The Journal of the American Society of Hematology 115, 4393–4402 (2010).

8. Rada, M., Barlev, N. & Macip, S. BTK: a two-faced effector in cancer and tumour suppression. Cell death & disease 9, 1–3 (2018).

9. Day, C.L., et al. PD-1 expression on HIV-specific T cells is associated with T-cell exhaustion and disease progression. Nature 443, 350–354 (2006).

10. Le, D.T., et al. PD-1 blockade in tumors with mismatch-repair deficiency. New England Journal of Medicine 372, 2509–2520 (2015).

11. Chamoto, K., Hatae, R. & Honjo, T. Current issues and perspectives in PD-1 blockade cancer immunotherapy. International journal of clinical oncology 25, 790–800 (2020).

12. Zeng, C., et al. Neutralization of SARS-CoV-2 Variants of Concern Harboring Q677H. Mbio 12, e02510–02521 (2021).

13. Collier, D.A., et al. Age-related immune response heterogeneity to SARS-CoV-2 vaccine BNT162b2. Nature 596, 417–422 (2021).

14. Shrock, E., et al. Viral epitope profiling of COVID-19 patients reveals cross-reactivity and correlates of severity. Science 370(2020).

15. Chemaitelly, H., et al. Waning of BNT162b2 vaccine protection against SARS-CoV-2 infection in Qatar. New England Journal of Medicine (2021).

16. Keehner, J., et al. Resurgence of SARS-CoV-2 infection in a highly vaccinated health system workforce. New England Journal of Medicine 385, 1330–1332 (2021).

17. Monin, L., et al. Safety and immunogenicity of one versus two doses of the COVID-19 vaccine BNT162b2 for patients with cancer: interim analysis of a prospective observational study. The Lancet Oncology (2021).

18. Shroff, R.T., et al. Immune responses to two and three doses of the BNT162b2 mRNA vaccine in adults with solid tumors. Nature Medicine, 1–10 (2021).

19. Greenberger, L.M., et al. Antibody response to SARS-CoV-2 vaccines in patients with hematologic malignancies. Cancer Cell 39, 1031–1033 (2021).

20. Greenberger, L.M., et al. Anti-spike antibody response to SARS-CoV-2 booster vaccination in patients with B cell-derived hematologic malignancies. Cancer cell (2021).

## Supplementary references

1. Zeng, C., et al. Neutralizing antibody against SARS-CoV-2 spike in COVID-19 patients, health care workers, and convalescent plasma donors. JCI insight 5(2020).

2. Mazurov, D., Ilinskaya, A., Heidecker, G., Lloyd, P. & Derse, D. Quantitative comparison of HTLV-1 and HIV-1 cell-to-cell infection with new replication dependent vectors. PLoS Pathog 6, e1000788 (2010).

3. Yu, J., et al. IFITM proteins restrict HIV-1 infection by antagonizing the envelope glycoprotein. Cell reports 13, 145–156 (2015).

4. Goerke, A.R., Loening, A.M., Gambhir, S.S. & Swartz, J.R. Cell-free metabolic engineering promotes high-level production of bioactive Gaussia princeps luciferase. Metabolic engineering 10, 187–200 (2008).

